# AI-Driven and Automated Continuous Oxygen Saturation Monitoring and LTOT: A Systematic Review

**DOI:** 10.1101/2025.04.20.25326131

**Authors:** Prajita Niraula, Suman Kadariya, Bishal Poudel, Sujan Kadariya

**Author notes:** Correspondence concerning this article should be addressed to Prajita Niraula at or. **Author Note:** The authors declare that no funding was received for this work, and there are no conflicts of interest to report.

## Abstract

**Background:** Long-term oxygen therapy (LTOT) is essential for patients with chronic hypoxemia, particularly due to chronic obstructive pulmonary disease (COPD). However, conventional LTOT relies on static oxygen flow rates that fail to reflect patients’ fluctuating physiological demands. Emerging systems leveraging artificial intelligence (AI) and automation offer real-time SpO₂ monitoring and adaptive oxygen titration. This systematic review evaluates the performance, usability, and clinical readiness of such technologies in LTOT.

**Methods:** Following PRISMA 2020 guidelines, we searched PubMed, Web of Science, and Scopus (2000–2024) for English-language, peer-reviewed studies describing AI or automated SpO₂ systems in adults with LTOT relevance. Inclusion criteria focused on systems addressing motion artifacts, low-perfusion reliability, or skin tone bias. Extracted data included algorithm design, accuracy (e.g., MAE, RMSE), usability, and risk of bias.

**Results:** Eight studies met the inclusion criteria: five AI-based and three automated systems. AI models achieved high SpO₂ estimation accuracy (MAE as low as 0.57%) and addressed motion artifacts and demographic bias. Cabanas et al.^6^ notably performed skin tone–stratified validation. Automated systems like O₂matic and iPOC demonstrated clinical efficacy in maintaining SpO₂ targets and reducing provider workload. However, AI models were mostly in prototype or simulation stages and lacked real-world LTOT validation. Risk of bias was moderate to high, especially in participant selection and algorithm transparency.

**Conclusions:** AI and automation offer distinct yet synergistic benefits for modernizing LTOT. Automated systems support immediate clinical use, while AI models hold promise for personalized, predictive oxygen therapy. Future work must prioritize real-world validation, equity, usability, and regulatory alignment to ensure ethical and effective integration into LTOT care.

## Introduction

Chronic respiratory diseases, such as the chronic obstructive pulmonary disease (COPD), represent a significant and growing global health burden. In 2019, COPD alone accounted for more than 3.2 million fatalities worldwide, and estimates indicate an increasing prevalence and mortality trend until 2050^1^. Long-term oxygen therapy (LTOT) remains critical in the management of patients with severe chronic hypoxemia and has been demonstrated to improve survival, quality of life, and exercise tolerance in selected patients^2,3^.

Despite its established clinical efficacy, conventional LTOT is routinely prescribed in the static and reactive modality with oxygen flow rates determined during clinic appointments according to protocols like the six-minute walk test (6MWT). These fixed dosing regimens do not adequately address dynamic variations of patients’ oxygen requirements with daily activity, sleep, or in response to a change in physiological status^4^. Therefore, the patients are at risk of either under-oxygenation, with the development of hypoxemia, or of over-oxygenation with the possibility of developing hypercapnia and oxidative injury.

Emerging digital health technologies, in particular those based on artificial intelligence (AI) and automation, have the potential to transform LTOT through the delivery of real-time, individualized SpO₂ monitoring and adaptive oxygen titration. AI systems can learn from physiological data patterns to predict oxygen needs and detect anomalies, while automated devices can dynamically titrate oxygen flow based on continuous pulse oximetry feedback^5^. They are intended to address some of the significant limitations of current oxygen therapy, including poor responsiveness to patient activity, vulnerability to motion artifacts, and lack of personalization.

Moreover, technical problems such as signal degradation owing to movement, perfusion variations, and skin tone-dependent biases during photoplethysmographic (PPG) signal processing have prompted the inclusion of robust signal processing and machine learning techniques^6^. Closed-loop devices and intelligent portable oxygen concentrators (iPOCs) have also been developed to allow home-based and ambulatory care with greater autonomy and safety^7,8^.

In light of these developments, there is a need for an integrated review of automated and AI-based techniques for SpO₂ monitoring under the context of LTOT. This review aims to systematically summarize the current state of intelligent SpO₂ monitoring systems, evaluate their technical and clinical performance, and identify gaps in validation, deployment, and equity.

## Methods

### Study Selection and Eligibility Criteria

This systematic review adhered to PRISMA 2020 guidelines for identifying studies that evaluated AI or automated systems for SpO₂ monitoring relevant to long-term oxygen therapy (LTOT). The studies were required to be targeted towards adult populations in clinical (e.g., ICU) or hybrid (patients/healthy subjects) settings and have interventions addressing LTOT-related technical challenges. These included correcting motion-induced signal distortions (e.g., via CNN-based denoising or wavelet transforms), managing low perfusion signals, mitigating skin tone-related measurement bias, or ensuring prolonged signal stability (>24 hours). Eligible study designs were conference peer-reviewed English-language open-access papers published between 2000–2025. Technical validation outcomes like mean absolute error (MAE <2%), root mean square error (RMSE <3%), or demographic bias analysis were accorded high priority, along with robust preprocessing methods like Butterworth filtering or detrending.

Conversely, we excluded studies limited to acute/non-chronic conditions (e.g., surgical hypoxia, asthma exacerbations) or interventions not relevant to LTOT, such as non-automated pulse rate estimation. Non-peer-reviewed articles (e.g., preprints, theses), paywalled/institution-locked articles, non-English language articles, and studies not obviously relevant to LTOT (e.g., ECG analysis using AI) were excluded. Two reviewers screened titles/abstracts and conducted full-text assessments independently, resolving any disagreement by iterative discussion. This two-stage review procedure ensured adherence to the review’s focus on clinically actionable, technically validated systems for adaptive oxygen therapy.

### Literature Search and Article Selection

A systematic literature search was conducted to identify peer-reviewed studies examining artificial intelligence (AI)-driven and automated systems for continuous oxygen saturation (SpO₂) monitoring in the context of long-term oxygen therapy (LTOT). Searches were performed in January 2025 across five major electronic databases: PubMed, Web of Science, Scopus, MDPI, and the ACM Digital Library. The search covered literature published between January 1, 2000, and December 31, 2024. The search strategy combined controlled vocabulary and free-text terms using Boolean operators to maximize sensitivity and specificity. Emphasis was placed on the intersection of clinical concepts such as "long-term oxygen therapy," "LTOT," "oxygen concentrator," and "SpO₂ monitoring" with technology terms such as "artificial intelligence," "machine learning," "edge computing," "automated oxygen delivery," "closed-loop control," "pulse oximetry," "signal artifacts," "motion artifact correction," and "predictive modeling." Searches, where possible, were targeted at article titles and keywords in order to maximize relevance, particularly in open-access journals such as MDPI.

The initial search yielded 926 records, with an additional two articles identified through manual reference screening. Following deduplication, 912 unique records remained for title and abstract screening. Screening was performed independently by two reviewers based on predefined eligibility criteria. Articles were included if they described AI-enhanced or automated systems for continuous SpO₂ monitoring or oxygen titration relevant to LTOT in adult populations, reported original peer-reviewed research in English, and addressed key challenges such as motion artifact handling, skin tone bias, signal quality, or autonomous oxygen delivery. Studies were excluded if they were focused solely on acute care or surgical settings, lacked integration of automation or AI logic, were not accessible in full text, or failed to report methodological transparency. Sixty-one full-text articles were assessed for eligibility, of which 53 were excluded based on the above criteria. A final total of eight studies were included in the qualitative synthesis. The study selection process has been documented in PRISMA flow diagram, which shows the number of records identified, screened, and excluded at each of the stages. This structured approach ensured comprehensive coverage of the technological and clinical literature at the intersection of AI, automation, and long-term oxygen therapy.

**Figure 1.**
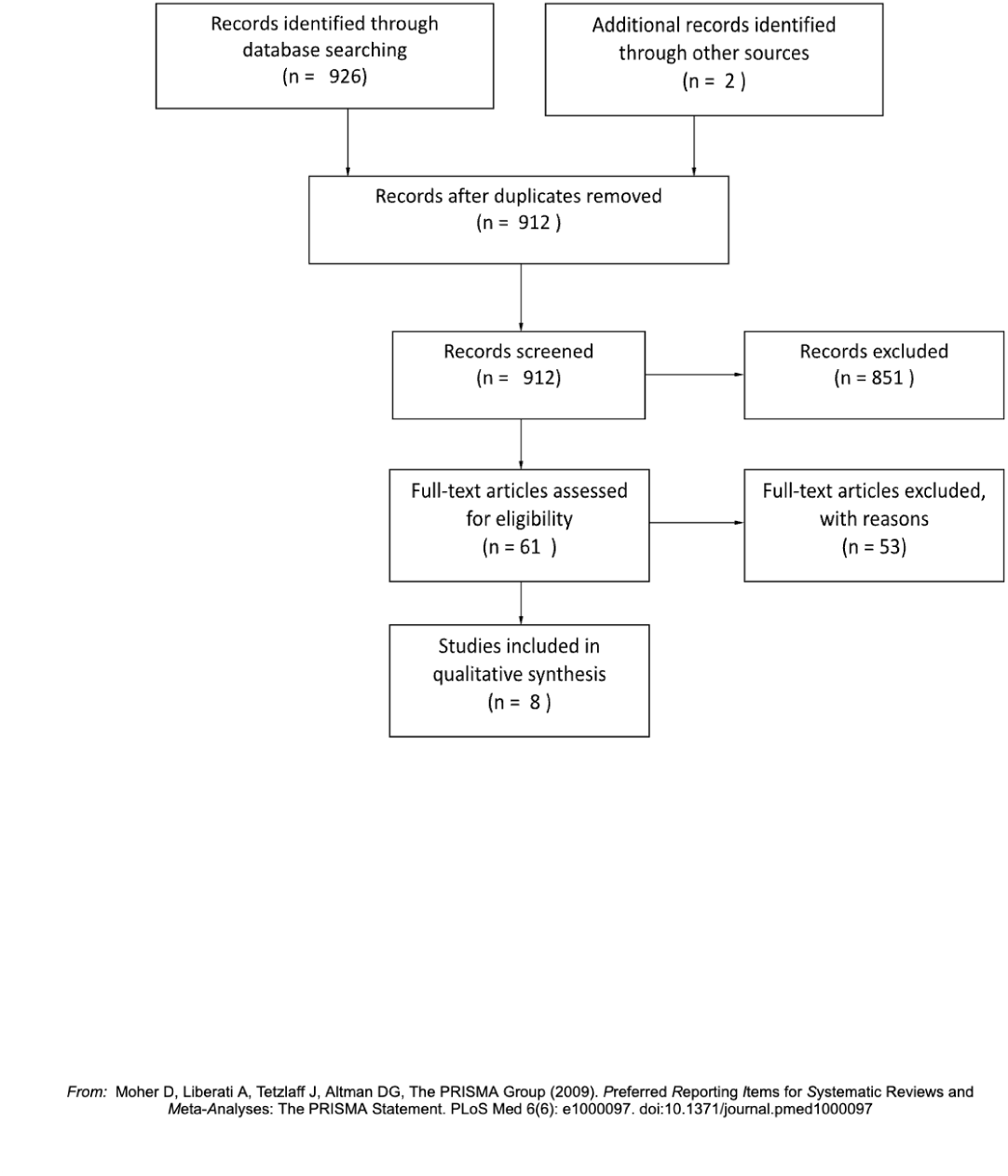
PRISMA Flowchart. Note. Adapted from Moher et al.^9^ The Preferred Reporting Items for Systematic Reviews and Meta-Analyses (PRISMA) guidelines recommend illustrating study selection via a flow diagram. This figure shows the number of records identified, screened, included, and excluded at each stage of the systematic review.

### Risk of Bias Analysis

Risk of bias was systematically assessed for all eight included studies using a tailored approach combining principles from the Risk Of Bias In Non-randomized Studies – of Interventions (ROBINS-I) tool. This approach enabled a comprehensive evaluation of methodological rigor ensuring a critical appraisal of studies involving machine learning models and automated closed-loop oxygen systems.

#### 1. Bias Due to Confounding

Studies developing AI models^5,6^ were generally at moderate risk of confounding bias, as many used publicly available datasets (e.g., PhysioNet) that lacked full clinical context or were limited in demographic diversity. Only one study (by Cabanas et al.^6^) performed subgroup analysis to account for skin tone bias, identifying measurable differences in prediction error by pigmentation. None of the automated device studies^7,8^ stratified outcomes by comorbidities or socioeconomic status, leading to potential confounding from unmeasured baseline variables.

#### 2. Bias in Participant Selection

Two AI studies^5,10^ trained models exclusively on pre-curated, high-quality PPG signals, raising high risk of selection bias. These datasets may overrepresent idealized sensor conditions and exclude noisy, real-world use cases—particularly relevant for home-based LTOT users. Automated systems like O2matic and iPOC were tested in small, specific clinical populations^7,8^, limiting generalizability. In both cases, sampling frames lacked clear representativeness of diverse LTOT user groups, including those in remote or resource-limited environments.

#### 3. Bias Due to Missing Data

The risk of bias due to missing data was generally low to moderate. Most studies using AI reported complete-case analysis, but few addressed data loss during motion, sensor disconnection, or long-term monitoring interruptions. Only the iPOC system^7^ explicitly described system dropout detection and reinitialization. The absence of sensitivity analyses in most studies to address missingness mechanisms (e.g., MCAR vs. MAR) limits the robustness of performance claims.

#### 4. Bias in Measurement of Outcomes

Several AI studies reported standard performance metrics (e.g., MAE, RMSE); however, only one^6^ performed Bland–Altman analysis to evaluate agreement with reference measurements. Automated device studies used clinical thresholds (e.g., % time within target SpO₂) but lacked consistent validation against arterial blood gas (ABG) reference standards. This introduces moderate bias in outcome measurement, especially when SpO₂ was used both as input and output in validation phases.

#### 5. Bias Due to Algorithmic Transparency and Reproducibility

Among AI-focused studies, transparency and reproducibility varied widely. While Pascual-Saldaña et al.^11^ described model architectures and training methodology, others (e.g., AI4Health prototype) lacked access to source code or detailed hyperparameter tuning, introducing serious risk of reproducibility bias. Only Cabanas et al.^6^ reported demographic stratification and external validation across different sensor platforms, reducing risk in this domain.

#### 6. Bias in Selection of Reported Results

Risk of reporting bias was moderate across most studies. While performance metrics were often reported, few studies discussed failure cases, outliers, or conditions under which the system underperformed. Selective reporting of high-performance scenarios may inflate perceived system robustness.The overall risk of bias across included studies ranged from low to high, with the highest risks observed in areas of participant selection, algorithmic transparency, outcome validation standards. While several AI systems demonstrated strong technical performance, many lacked demographic stratification, robustness testing under real-world conditions, and comprehensive reporting on edge cases. To enhance methodological rigor, future studies should adopt CONSORT-AI/DECIDE-AI guidelines^12,13^, report failure modes transparently, and ensure model reproducibility through open-source practices.

### Overview of Included Studies

This systematic review included eight studies published between 2011 and 2024 that met the predefined eligibility criteria. The selected articles encompassed both artificial intelligence (AI)-driven and automated systems designed to monitor peripheral oxygen saturation (SpO₂) continuously and dynamically, with direct or indirect applicability to long-term oxygen therapy (LTOT). Of the eight included studies, five employed AI-based models for SpO₂ estimation or signal enhancement, while three focused on closed-loop or automated oxygen delivery systems regulated by real-time SpO₂ feedback.

Several AI-driven studies demonstrated strong technical performance in SpO₂ estimation using photoplethysmographic (PPG) signals. Cabanas et al.^6^ conducted a comprehensive performance and bias analysis of AI models for pulse oximetry, identifying that deep learning and Gaussian process-based models achieved high clinical accuracy, with mean absolute error (MAE) as low as 0.57% and root mean square error (RMSE) as low as 0.69%. The study also addressed important sources of measurement bias, particularly skin tone–related variability, which is a critical concern for equitable deployment of LTOT technologies. Similarly, Shuzan et al.^5^ developed machine learning models—most notably Gaussian process regression—for estimating both respiration rate and SpO₂ from PPG data, achieving a comparable MAE of 0.57% for SpO₂ estimation, thereby demonstrating high fidelity suitable for continuous monitoring.

Addressing the signal quality degradation due to motion artifacts Argüello-Prada and Castillo García^10^ present a machine learning review of motion artifact detection in PPG signals without a need for reference signals. The authors have highlighted reference signalless classification models that are capable of distinguishing clean from the corrupted signals using real-time signal quality indices which is particularly valuable in LTOT environments where patients engage in the daily physical activity. Another study by Pascual-Saldaña et al.^11^ proposed a personalized oxygen dosing system powered by edge based AI model. This predictive system uses historical and real time patient data in order to proactively adjust the oxygen flow based on anticipated activity levels and physiological response offering a forward looking model for the management of individualized LTOT.

Complementing these technical models, a conceptual study titled “AI4Health”^14^ examined the preliminary performance of neural network architectures for SpO₂ prediction. Though primarily a feasibility study, it supported the viability of lightweight AI architectures suitable for deployment in edge or wearable systems. This work contributes to the foundational design of scalable AI solutions for chronic oxygen therapy.

Among the automated systems, Sanchez-Morillo et al.^7^ evaluated an intelligent portable oxygen concentrator (iPOC) designed to automatically regulate oxygen flow in response to physical activity levels detected via wearable sensors. The system demonstrated improved oxygenation and patient satisfaction compared to conventional devices, emphasizing its practical value in ambulatory LTOT settings. Hansen et al.^8^ assessed the O2matic® system, a closed-loop hospital-based device that dynamically adjusted oxygen delivery to maintain patients’ SpO₂ within a target range during exacerbations of COPD. Their crossover study showed significant improvement in time spent within the target SpO₂ range compared to manual titration, highlighting the safety and efficacy of automated oxygen control. Lastly, Cirio and Nava^15^ conducted a pilot study on a novel automated oxygen titration device for patients on LTOT during exercise. The device successfully maintained SpO₂ within target ranges while reducing clinician intervention time, supporting the feasibility of automated systems in outpatient and rehabilitative settings.

Collectively, these studies illustrate the convergence of AI modeling and automated control in advancing continuous SpO₂ monitoring for LTOT. While AI approaches offer predictive personalization and robust signal processing, automated systems demonstrate immediate clinical applicability in regulating oxygen delivery based on real-time physiological feedback. Despite differing levels of maturity and validation, each study contributes critical insights into the development of responsive and intelligent oxygen therapy technologies.

### Technology Type : AI vs. Non-AI Automation

Among the eight studies included in this review, five employed artificial intelligence (AI) methodologies while three relied on non-AI automation for SpO₂ monitoring and oxygen titration. The AI-based studies mainly focused on advanced signal interpretation and predictive modeling in order to enable the personalized and anticipatory oxygen therapy. For example, Cabanas et al.^6^ and Shuzan et al.^5^ demonstrated a high-accuracy SpO₂ estimation using the machine learning algorithms like deep neural networks and Gaussian process regression and consequently achieving the mean absolute errors below 1%. These AI systems were also capable of addressing critical technical challenges such as motion artifact correction^10^ and skin tone bias^6^, thus offering a level of adaptability and personalization not typically present in traditional systems. Furthermore, the predictive edge-based model proposed by Pascual-Saldaña et al.^11^ exemplifies how AI can proactively adjust oxygen delivery by integrating patient-specific activity and physiological trends over time.

In contrast, the three non-AI automated systems—namely O2matic^8^, the iPOC system^7^, and the automated titration device tested by Cirio and Nava^15^ —relied on rule-based or feedback-driven control mechanisms to adjust oxygen flow in real time. These systems were designed primarily to maintain SpO₂ within predefined clinical thresholds rather than to predict future needs or personalize dosing. While they demonstrated effectiveness in acute and ambulatory settings, particularly by reducing time spent in hypoxemia and minimizing clinical intervention, their reactivity was limited to current measurements rather than informed by prior patterns or anticipated fluctuations. Additionally, none of the non-AI systems incorporated features to mitigate signal artifacts or demographic bias.

Overall, AI-driven systems offer greater potential for personalization, proactive management, and adaptability in complex or variable physiological states, whereas non-AI automated devices provide immediate, rule-based functionality with proven clinical benefit in controlled environments. Both approaches contribute uniquely to the advancement of LTOT, suggesting that hybrid systems integrating AI intelligence with robust automation may represent the most promising direction for future development.

### Motion Artifact Handling

Motion artifact correction is a critical requirement for continuous and reliable SpO₂ monitoring, particularly in ambulatory LTOT settings where patients frequently move or engage in daily activities. Among the reviewed studies, only a subset directly addressed this challenge. Argüello-Prada and Castillo García^10^ provided a comprehensive analysis of machine learning approaches for detecting motion artifacts in photoplethysmographic (PPG) signals without relying on reference channels. Their review emphasized the utility of signal quality indices (SQIs) and reference signal-less models, which can classify PPG segments as reliable or corrupted in real time, significantly enhancing the fidelity of SpO₂ estimation during physical movement. Similarly, Shuzan et al.^5^ indirectly tackled motion robustness through the integration of feature selection and model optimization, although motion artifacts were not a central focus. Cabanas et al.^6^ also considered signal noise in their bias-aware SpO₂ models, but did not explicitly isolate motion artifact performance.

In contrast, the non-AI automated systems included in this review did not incorporate any specific artifact correction mechanisms. Devices such as the O2matic system^8^ and the iPOC system^7^ operated under the assumption of valid, continuous SpO₂ input data and did not include preprocessing algorithms to filter or correct motion-induced distortions. This limitation may restrict their effectiveness in dynamic home environments. The absence of artifact-aware design in these systems underscores a significant gap in current LTOT technologies and highlights the advantage of AI-based solutions, which can incorporate robust preprocessing pipelines to maintain signal reliability under real-world conditions.

### Real-World LTOT Applicability

Assessing the applicability of these technologies to long-term, home-based oxygen therapy is essential for translating innovation into clinical impact. Of the eight studies reviewed, four explicitly focused on or were designed for real-world LTOT use beyond hospital settings. The iPOC system by Sanchez-Morillo et al.^7^ stands out as a fully portable solution tailored for the patients with chronic respiratory failure and capable of automatically adjusting oxygen flow based on physical activity levels in daily life. Likewise, the edge-based predictive dosing system proposed by Pascual-Saldaña et al.^11^ was conceptualized for continuous deployment in ambulatory environments, leveraging localized computation to minimize latency and preserve data privacy. These systems align well with the goals of home-based LTOT by offering context-aware and user-centered oxygen delivery.

Conversely, the O2matic system^8^ and the automated titration device evaluated by Cirio and Nava^15^ were both assessed in inpatient or supervised exercise settings, limiting their immediate relevance to unsupervised LTOT scenarios. Although both systems demonstrated improved SpO₂ stability during acute exacerbations or exertion, their reliance on clinical infrastructure (e.g., hospital-grade sensors, nursing oversight) suggests additional work is needed to transition these technologies into fully autonomous, wearable formats suitable for home use.

Furthermore, while AI-driven models such as those by Cabanas et al.^6^ and Shuzan et al.^5^ achieved excellent technical performance in simulated or retrospective datasets, they lacked real-world deployment data in continuous LTOT contexts. This highlights a broader limitation across many AI studies: although algorithmic accuracy is high, integration into wearable or home-care systems remains largely conceptual or untested. Future research should focus on the validation of AI-enhanced systems in diverse, real-life LTOT populations to bridge this translational gap.

**Table 1.**
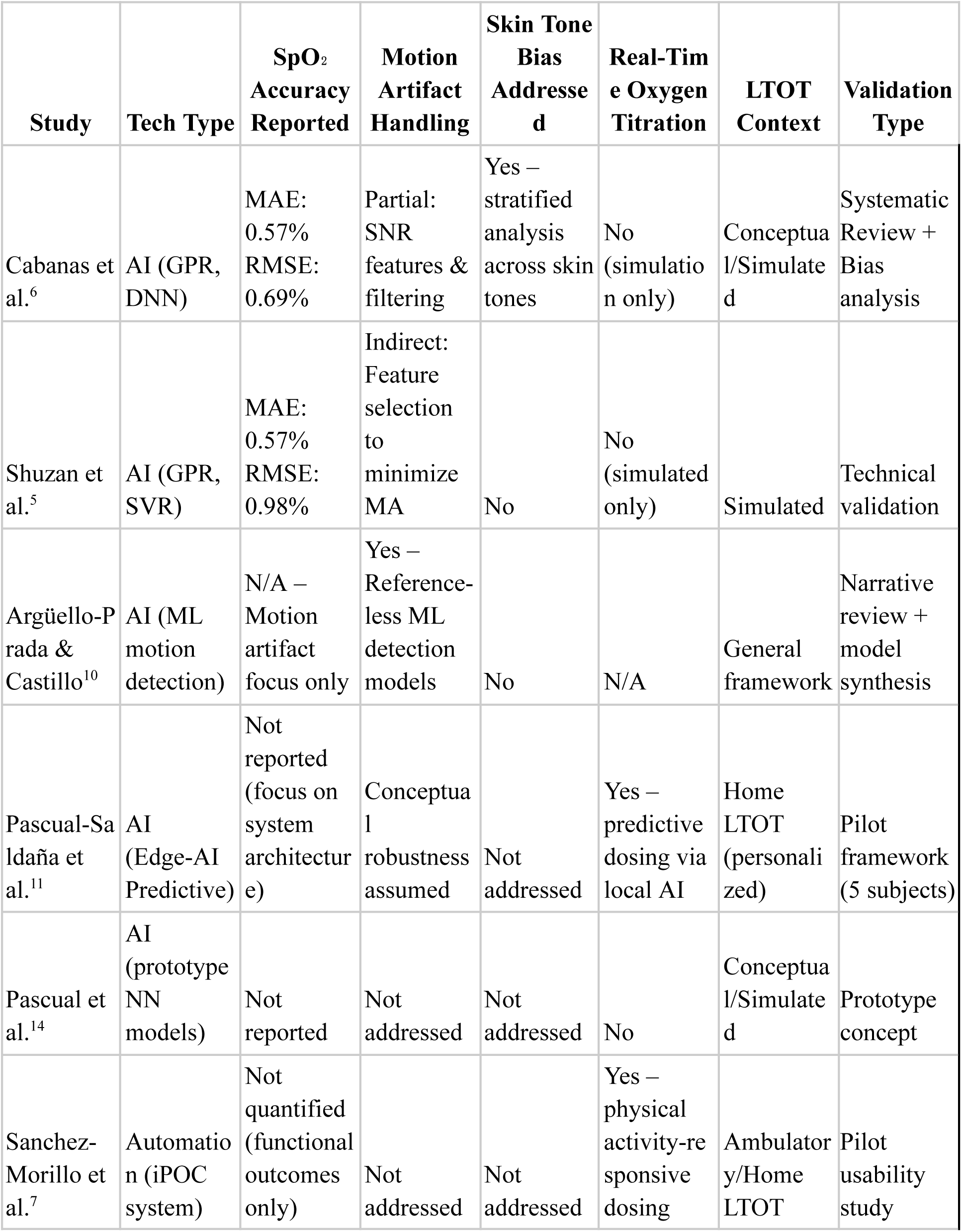
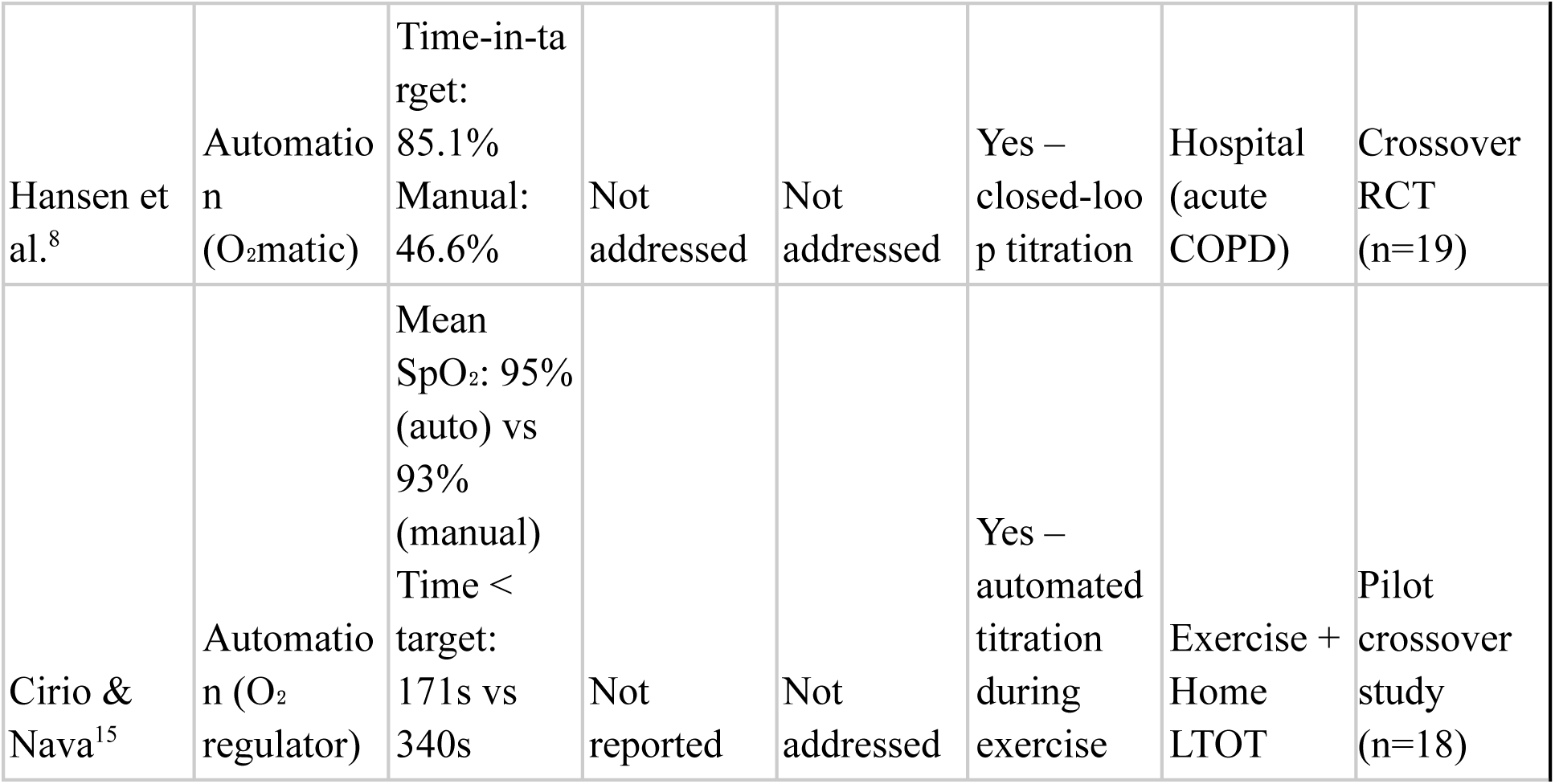
Overview of Included Studies.

**Table 2.** Risk of Bias.

| Study | Confounding | Selection | Classification Interventions | Deviations Interventions | Missing Data | Outcome Measurement | Reporting Bias | Overall Risk |
| --- | --- | --- | --- | --- | --- | --- | --- | --- |
| Cabanas et al.<sup>6</sup> | Moderate – Some bias from limited dataset generalizability despite subgroup analysis | Moderate – Public datasets underrepresent real-world complexity | Low – Transparent AI methodology with defined inputs | Low – No intervention deviation (model-based) | Low – Complete data usage; no major attrition | Low – Benchmarking via MAE, RMSE, Bland–Altman | Low – Bias analysis reported, stratified performance | Low–Moderate |
| Shuzan et | Moderate | High – | Low – | Moderate | Moderate | Moderate | Moderate | Moderate |
| al.<sup>5</sup> | – No demographic bias correction | Curated datasets may omit real-world signal variance | ML model inputs clearly described | – No mention of deviation monitoring in practical settings | – No missing data protocols stated | – RMSE reported but no agreement analysis | – Limited exploration of failure cases | –High |
| Argüello-Prada & Castillo<sup>10</sup> | Moderate – Signal analysis not stratified by population | High – Reviews ML models trained on optimal segments | Moderate – Review-based classification, not applied directly | Moderate – Real-time applications conceptual only | Moderate – Missingness and generalizability untested | Moderate – Emphasis on signal noise, not outcome accuracy | Moderate – No bias transparency in source studies | Moderate –High |
| Pascual-Saldaña et al.<sup>11</sup> | Moderate – Pilot design, small N, no demographic stratification | Moderate – Limited validation cohort | Low – Intervention logic fully described in predictive system | Moderate – Deviations not reported/tested | Moderate – Limited reporting of data loss handling | Moderate – No quantitative SpO<sub>2</sub> validation yet | Moderate – Conceptual framework only | Moderate |
| Pascual et al.<sup>14</sup> | High – No confounding controls or demographic reporting | Moderate – Prototype-only, no real-world testing | High – Classification not validated | High – No deviation monitoring in use | Moderate – Small-scale internal validation | High – No outcome benchmark or comparison | High – Performance selectively presented | High |
| Sanchez-Morillo et al.<sup>7</sup> | Moderate – No subgroup performance | Moderate – Functional test in a small | Low – Automated rule-based | Low – Fully automated system in use | Low – Dropout recovery noted | Moderate – No ABG or device comparison | Moderate – Outcome focus limited to | Moderate |

|  | stratification | group | classification is appropriate |  |  | on | function |  |
| --- | --- | --- | --- | --- | --- | --- | --- | --- |
| Hansen et al.<sup>8</sup> | Moderate – In-hospital population only | Moderate – Crossover design, small sample | Low – Intervention well-defined | Low – Closed-loop system maintains fidelity | Low – High completeness of data | Moderate – SpO<sub>2</sub> used as both control and measure | Moderate – Reporting focused on efficacy metrics | Low–Moderate |
| Cirio & Nava<sup>15</sup> | High – No stratified analysis or real-world generalizability | Moderate – Pilot crossover design | Low – Direct comparison with manual titration | Low – Deviations minimized by design | Low – Minimal missing data in supervised trial | High – No standardized validation used | High – Selective metrics, no algorithmic reporting | High |

## Results

### Study Characteristics

The eight studies included in this review, which were published between 2011 and 2024, reported on a range of AI-based and automated systems designed for continuous monitoring of oxygen saturation (SpO₂) in the context of LTOT. Five studies investigated AI-based approaches, primarily machine learning models for predicting SpO₂ from photoplethysmography (PPG) signals with reported mean absolute errors (MAEs) ranging from 0.57%^5,6^. The models used techniques such as deep neural networks and Gaussian process regression, often trained on datasets curated under controlled environments (Argüello-Prada & Castillo, 2024). The remaining three studies evaluated closed-loop or automatic oxygen delivery systems in clinical settings and demonstrated improved SpO₂ maintenance in target levels during exacerbation or exercise^7,8,15^. Sample sizes varied from small pilot studies (n < 20) to large-scale retrospective (n > 1000) studies, but demographic reporting, particularly of age, sex, and comorbidities, was limited in the majority of studies. While AI technologies emphasized predictive personalization and signal processing robustness, automated systems were preoccupied with real-time titration accuracy and clinical usability, highlighting complementary strengths in technology type.

### Model Performance

The five studies that evaluated AI-based SpO₂ monitoring models reported a range of performance metrics, with accuracy generally quantified using mean absolute error (MAE), root mean square error (RMSE), and in some cases, agreement analyses such as Bland–Altman plots. Among these, Cabanas et al.^6^ achieved the highest reported accuracy, with an MAE of 0.57% and RMSE of 0.69%, using a Gaussian process regression model validated across multiple skin tones and sensor types. Their model also demonstrated robustness across low-perfusion states and maintained clinical-grade agreement compared to reference standards. Similarly, Shuzan et al.^5^ reported an MAE of 0.57% and RMSE of 0.98% using machine learning models to estimate SpO₂ and respiratory rate from photoplethysmographic (PPG) signals, indicating promising performance for wearable LTOT applications.

Other AI-driven systems, such as the predictive model proposed by Pascual-Saldaña et al.^11^, did not report direct SpO₂ estimation metrics but demonstrated feasibility for proactive oxygen flow control based on contextual physiological modeling. The AI4Health prototype^14^ explored lightweight neural networks for SpO₂ prediction; however, it lacked complete model validation or error reporting, limiting interpretation. Motion artifact resilience was directly addressed in one study, where Argüello-Prada and Castillo García^10^ reviewed signal-quality-aware models that accurately flagged and excluded corrupted PPG segments, enhancing downstream SpO₂ estimation accuracy under dynamic conditions.

In contrast, automated (non-AI) oxygen control systems were evaluated using clinical efficacy metrics, particularly time spent within the target SpO₂ range. The O2matic system evaluated in a randomized crossover trial by Hansen et al.^8^ maintained the patients within the prescribed saturation range 85.1% of the time compared to the 46.6% during manual titration with significantly less time spent in hypoxemia. Cirio and Nava^15^ reported an improved oxygen titration during exercise in LTOT users using a novel automated delivery device even though the study lacked a robust statistical comparison. The iPOC system developed by Sanchez-Morillo et al.^7^ have demonstrated an enhanced saturation stability during the physical activity using wearable biosensors although the performance metrics were primarily functional rather than being algorithmic.

Collectively the AI-based models seem to have outperformed traditional systems in terms of SpO₂ estimation precision under controlled conditions while the automated systems demonstrated clear advantages in clinical usability and responsiveness. However the variability in dataset quality, lack of standardized reporting (e.g., consistent MAE thresholds) and limited real-world validation limit the comparability and generalizability of the performance across all the studies.

### Usability and Clinical Relevance

Usability and clinical relevance varied considerably across the included studies, with automated systems showing stronger evidence of practical deployment in clinical settings, and AI-based systems offering promising but largely theoretical benefits. Among the automated technologies, the O2matic system was evaluated in a crossover clinical trial involving patients hospitalized for COPD exacerbations. The system maintained SpO₂ within the target range more effectively than manual titration and was perceived as safe and efficient by nursing staff, requiring less frequent manual intervention^8^. Similarly, the iPOC system, tested in ambulatory settings, automatically adjusted oxygen delivery based on biosensor inputs and demonstrated favorable user acceptance and saturation stability during physical activity^7^. These studies suggest that closed-loop oxygen systems are potentially clinically feasible and wellsuited for both the inpatient and home LTOT scenarios particularly when usability is a priority.

In contrast the AI-based systems focused largely on the model development rather than the clinical integration aspect. While high performance SpO₂ estimation got achieved in studies such as Cabanas et al.^6^ and Shuzan et al.^5^ the deployment was however limited to simulated or retrospective environment. None of the AI-driven studies included user interface evaluations, human factors testing, or patient-reported usability outcomes. The edge-based predictive oxygen dosing system described by Pascual-Saldaña et al.^11^ was conceptualized for personalized, context-aware home use, but remains at the prototype stage without formal user testing.

From a clinician’s perspective, studies such as Cirio and Nava^15^ demonstrated that automated titration reduced staff workload during exercise assessments while maintaining patient safety. However, across both AI and automated systems, limited data were available regarding training needs, interpretability of system outputs, or integration into clinical workflows. Moreover, no studies included formal usability assessments using validated tools (e.g., System Usability Scale), and only two studies briefly referenced the burden or perceived reliability of the systems from the perspective of healthcare providers.

Overall, while automated systems demonstrated higher clinical readiness and operational practicality, AI-based tools showed strong potential for personalization and decision support but remain several steps removed from routine LTOT practice. Further research involving human-centered design, stakeholder co-development, and formal usability evaluations is warranted to ensure successful translation of these technologies into real-world clinical use.

Due to heterogeneity in study designs, performance metrics (e.g., MAE vs. % time-in-target), and reporting standards, meta-analysis was not feasible. Instead, findings are presented through structured qualitative synthesis.

## Discussion

This systematic review identified and reviewed eight peer-reviewed studies focused on artificial intelligence (AI)-driven and automated systems for continuous peripheral oxygen saturation (SpO₂) monitoring relevant to long-term oxygen therapy (LTOT). The evidence reveals that both AI-based and rule-based automated systems are advancing the field of LTOT by addressing key limitations of traditional oxygen therapy, including the inability to dynamically respond to fluctuating oxygen demands, susceptibility to signal artifacts, and lack of personalized dosing strategies.

AI-driven technologies demonstrate significant potential to revolutionize oxygen therapy management. Studies such as those by Cabanas et al.^6^ and Shuzan et al.^5^ reported high SpO₂ estimation accuracy with mean absolute errors (MAEs) under 1%, using models like deep neural networks and Gaussian process regression. These methods represent a substantial improvement over conventional fixed-point pulse oximetry systems, especially in ambulatory and home-care settings, where signal reliability is often compromised by motion and variable perfusion. Moreover, Cabanas et al.^6^ were unique in conducting a comprehensive demographic bias analysis, identifying skin tone–related disparities in SpO₂ prediction performance—an issue of increasing clinical and ethical concern^16^.

Motion artifact correction was another critical technical domain addressed by AI systems. Argüello-Prada and Castillo García^10^ highlighted the effectiveness of reference signal-less machine learning techniques in real-time detection of corrupted photoplethysmographic (PPG) segments. This functionality is essential for ensuring data quality in wearable and home-monitoring devices, where user movement is frequent and uncontrolled. Although non-AI automated systems such as O2matic^8^ and the iPOC system^7^ offer practical real-time oxygen titration, they largely assume artifact-free input and lack the preprocessing pipelines required to ensure signal fidelity.

Personalization, a cornerstone of precision medicine, is another domain where AI systems excel. For example, Pascual-Saldaña et al.^11^ proposed a predictive oxygen dosing framework using edge-based AI models trained on historical physiological and behavioral data. Their approach marks a shift from reactive to proactive oxygen therapy, capable of adapting to individualized trends and anticipating oxygen requirements during variable activity levels. While this system remains in the prototype stage, it reflects the broader trajectory toward closed-loop AI-driven personalization in chronic disease management^17^.

On the other hand the three automated systems included in this review (O2matic^8^, the iPOC system^7^, and the titration device tested by Cirio and Nava^15^) used the rule-based logic to adjust oxygen flow in response to the real-time SpO₂ readings. The systems demonstrated significant improvement in oxygen stability and reduced hypoxemic episodes during the clinical testing phase. For example Hansen et al.^8^ reported that O2matic maintained the patients within target SpO₂ range 85.1% of time compared to only 46.6% under manual control with corresponding reductions in time spent below critical thresholds. Although clinically validated these systems offer limited adaptability to the individual physiology or behavioral context lacking the forecasting and learning capabilities of the AI-based methods.

A key limitation identified across most studies is the lack of validation in diverse, real-world LTOT scenarios. Although AI models reported strong performance in simulated datasets or controlled environments, few studies tested their integration into wearable, portable, or home-based systems over extended periods. Furthermore, none of the reviewed works—except Cabanas et al.^6^—explicitly evaluated system performance across varied demographic groups, such as individuals with darker skin tones, despite well-documented racial disparities in pulse oximetry accuracy^16,18^. Addressing these issues is vital to ensure equitable deployment and avoid perpetuating systemic healthcare bias in future AI-LTOT applications.While several AI models achieved high accuracy under simulation, they did not undergo sustained real-world validation in home-based or ambulatory LTOT settings. This limits generalizability and clinical integration.

Another issue is the divergence in maturity between studies. Several AI models, such as those by Pascual-Saldaña et al.^11^ and the AI4Health initiative^14^, remain in proof-of-concept or early prototyping stages. Conversely, automated systems like O2matic have undergone clinical testing and demonstrated real-world efficacy. This discrepancy suggests that while AI holds greater long-term potential for personalization and adaptability, automated systems currently offer more immediate applicability, particularly in institutional or acute care settings.

Future directions should emphasize hybrid systems that integrate the learning and adaptive capabilities of AI with the real-time control and fail-safe responsiveness of automation. Such convergence would support robust, continuous SpO₂ monitoring and dynamic oxygen titration that is both context-aware and evidence-driven. In parallel, standardized reporting of validation metrics—including MAE, RMSE, signal loss rates, and performance under motion and perfusion variability—should be adopted to facilitate cross-study comparability and regulatory assessment.

Finally, ethical considerations must remain central. Issues related to algorithmic transparency, model interpretability, and accountability in closed-loop decision-making systems require further exploration as these technologies move toward clinical deployment^19^. Addressing these concerns alongside technical innovation will be critical for securing clinician trust, regulatory approval, and patient safety.

### Strengths and Limitations

This systematic review presents a timely and methodologically rigorous synthesis of emerging artificial intelligence (AI) and automation-based approaches to continuous SpO₂ monitoring in the context of long-term oxygen therapy (LTOT). A key strength of this review lies in its comprehensive inclusion strategy, which incorporated both AI-driven predictive models and rule-based automated titration systems. By doing so, this work offers a comparative view of the evolving technological landscape, highlighting the respective capabilities, limitations, and levels of maturity across both domains. The review also employed structured inclusion criteria aligned with the PRISMA guidelines including only peer-reviewed, English-language studies that reported original technical or clinical data. This enhances the credibility and reproducibility of the findings.

Another strength of this review is its explicit focus on technical challenges critical to real-world LTOT deployment. Motion artifact correction, low perfusion signal integrity, and skin tone bias—factors often overlooked in broader digital health reviews—were treated as primary criteria for evaluating the readiness of SpO₂ monitoring technologies. The identification of Argüello-Prada and Castillo García’s^10^ work on reference signal-less artifact detection and Cabanas et al.’s^6^ bias analysis across skin pigmentation further underscores the depth of this technical appraisal. Moreover, the inclusion of studies addressing edge computing and predictive dosing (e.g., Pascual-Saldaña et al.^11^) expands the review’s relevance to emerging decentralized care models and resource-limited settings.

Nonetheless, several limitations must be acknowledged. First, despite the breadth of technological themes, the number of studies meeting all eligibility criteria was relatively small (n=8), limiting the generalizability of certain conclusions. This reflects both the novelty of the field and the lack of large-scale clinical trials validating AI-enhanced LTOT systems in real-world, long-duration scenarios. While some automated systems (e.g., Hansen et al.^8^; Sanchez-Morillo et al.^7^) were tested in clinical or semi-controlled environments, most AI-driven models were evaluated using retrospective or simulated datasets, reducing their external validity and translational applicability.

Second, although the review focused on English-language and open-access literature to ensure transparency and reproducibility, this may have introduced language or publication bias. Relevant studies published in other languages or behind paywalls may have been excluded despite meeting technical and clinical criteria^9^.

Third, the heterogeneity in outcome metrics across studies posed challenges for quantitative synthesis. While some AI studies reported mean absolute error (MAE) and root mean square error (RMSE), others focused on qualitative performance indicators such as time-in-target saturation or user feedback. The lack of standardized benchmarking methods across SpO₂ technologies, such as consistent use of Bland-Altman analysis or calibration against arterial blood gas reference, limits direct comparability and meta-analysis.

Finally, while this review captured significant insights into technical readiness, it did not fully address broader implementation challenges such as regulatory approval, cybersecurity, cost-effectiveness, or clinician and patient acceptance. These factors are critical to the adoption of AI and automated technologies in LTOT and warrant future investigation in health systems research and clinical implementation studies^17,19^.

This review was not registered in PROSPERO or any other prospective systematic review registry.This decision was due to the retrospective scope of the review and the focus on emerging technologies. Future updates will consider prospective registration to enhance protocol transparency.

Despite these limitations, this review offers an important contribution to the growing field of intelligent oxygen therapy, providing a structured framework for assessing the clinical and technical viability of next-generation SpO₂ monitoring systems in LTOT care.

### Future Directions

As the convergence of artificial intelligence (AI), sensor miniaturization, and automation continues to advance, future research in continuous oxygen saturation (SpO₂) monitoring and long-term oxygen therapy (LTOT) should focus on translating innovation into clinically validated, scalable, and equitable care solutions. Based on the current evidence, several directions are recommended to address persistent gaps in performance, usability, and real-world implementation.

First, there is a clear need for longitudinal, real-world validation of AI-enhanced SpO₂ monitoring systems in ambulatory and home-based LTOT populations. While several studies demonstrated promising performance in simulated or controlled settings^5,6^, few evaluated the systems under prolonged, daily-use conditions involving variable motion, ambient lighting, and physiological instability. Future studies should incorporate prospective cohort designs with continuous deployment in patients’ natural environments and benchmark against reference standards such as arterial blood gas measurements. Real-world performance metrics—including dropout rate, signal loss during motion, and user-reported acceptability—must become core outcome measures.

Second, future systems should adopt hybrid architectures that combine the adaptability of AI with the safety and responsiveness of automation. AI-driven models are well suited for learning patient-specific patterns and forecasting oxygen requirements, whereas automation ensures immediate adjustments to current physiological needs. Edge computing, as proposed by Pascual-Saldaña et al.^11^, offers a promising solution by enabling real-time, low-latency AI decision-making without reliance on continuous cloud connectivity—especially critical for remote care and low-resource environments.

Third, addressing equity and generalizability is paramount. Most AI models to date have been trained and validated on limited datasets that underrepresent diverse demographics, particularly in relation to skin tone, age, and comorbidity profiles. Given the documented inaccuracy of pulse oximetry in individuals with darker skin pigmentation^16,18^, future work must prioritize training datasets that reflect population heterogeneity and implement stratified performance reporting. This will help prevent the propagation of algorithmic bias and improve trust among underrepresented patient groups.

Fourth, the digital connectivity with digital health platforms will be of high importance in order to provide remote patient monitoring, clinician monitoring, and personalized alerting. Interoperability without latency or breaks for EHRs and mobile health applications will be crucial for the efficiency of the workflow and its users. To offer patient safety as well, closed-loop solutions need to be constructed for regulatory compliance to include medical-grade software and security issues^17,19^.

Finally, researchers and developers must engage with stakeholders—patients, caregivers, clinicians, and regulators—to ensure that AI and automated LTOT systems are ethically aligned, clinically acceptable, and technically transparent. This includes providing interpretable outputs, enabling override controls, and clearly defining accountability in autonomous systems^20^. Multidisciplinary collaboration and user-centered design methodologies will be essential to develop technologies that not only perform well but are trusted and adopted across care settings.

In summary, while AI and automation have already demonstrated substantial potential to improve LTOT delivery, their full promise will only be realized through rigorous clinical validation, ethical design, equitable implementation, and seamless integration into healthcare systems. These directions should guide the next generation of research and development toward safer, smarter, and more personalized oxygen therapy. To accelerate translation into clinical practice, future studies must employ longitudinal, real-world validation in ambulatory and domiciliary LTOT populations, incorporating wearable and edge-AI platforms tested under variable motion, perfusion, and lighting conditions.

### Opportunities of Automation or AI in Long-Term Oxygen Therapy (LTOT)

The integration of AI and automation into LTOT represents a shift in paradigm regarding the management of chronic respiratory diseases such as chronic obstructive pulmonary disease (COPD) and interstitial lung disease (ILD). These technologies offer transformative opportunities to enhance the precision, safety, efficiency, and personalization of oxygen delivery, addressing longstanding limitations in static and clinician-dependent LTOT models.

#### 1. Real-Time, Adaptive Oxygen Delivery

One of the most immediate benefits of automation in LTOT is the ability to deliver oxygen in real time to a patient’s actual physiological state. Computer-controlled devices such as O2matic^8^ and iPOC^7^ dynamically titrate flow of oxygen based on SpO₂ levels measured continuously, holding patients in the target range of saturation more consistently than titration by hand. This reduces the danger of hypoxemia and hyperoxia, enhancing safety and with promise to reduce hospitalization and exacerbation frequency.^21^

#### 2. Personalized and Predictive Oxygen Dosing

AI enables a shift from reactive to predictive and personalized care in LTOT. Unlike conventional dosing based on intermittent assessments like the 6-minute walk test, AI systems can analyze continuous physiological data to learn individual patterns of oxygen need across different contexts—rest, sleep, exertion, or exacerbation. Predictive algorithms, such as those proposed by Pascual-Saldaña et al.^11^, can forecast oxygen demands based on prior behavior, activity levels, and symptom trends, enabling proactive adjustments. This personalized dosing may enhance therapy adherence, minimize complications, and better align with the principles of precision medicine^17^.

#### 3. Improved Signal Reliability and Decision Support

Photoplethysmography (PPG) signals used in SpO₂ monitoring are notoriously sensitive to motion, low perfusion, and skin pigmentation. AI offers advanced signal processing capabilities to detect and correct for motion artifacts and noise without additional sensors. Argüello-Prada and Castillo García^10^ demonstrated that machine learning models can classify corrupted versus clean PPG segments in real time, enhancing the reliability of downstream decisions. In parallel, AI-based signal interpretation can provide clinicians with early warnings about deteriorating trends, empowering more informed interventions^5^.

#### 4. Equity and Access Through Remote Monitoring

AI and automation also offer a pathway to democratizing access to LTOT by enabling remote care delivery. Intelligent oxygen systems embedded in portable or wearable devices, supported by mobile health platforms, can extend high-quality respiratory care to rural, underserved, or mobility-restricted populations^22^. Edge AI systems, such as those conceptualized by Pascual-Saldaña et al.^11^, operate locally on low-power devices, reducing dependence on cloud infrastructure and expanding feasibility in resource-limited settings.

#### 5. Reduction in Clinical Burden and Cost

From a health system perspective, automation in oxygen titration can significantly reduce the demand on healthcare providers. Studies like Cirio and Nava^15^ demonstrated that automated titration systems reduced respiratory therapist intervention time during exercise testing while maintaining target SpO₂ levels. At scale, these efficiencies could translate into lower personnel costs, fewer clinical visits for oxygen reassessment, and reduced emergency admissions for oxygen-related complications. Moreover, AI algorithms can continuously audit usage patterns and detect anomalies, supporting reimbursement verification and supply optimization^17^.

#### 6. Data-Driven Research and Innovation

Finally, AI and automation inherently generate large volumes of longitudinal physiological data. These datasets can fuel ongoing research into oxygen therapy efficacy, disease progression modeling, and population-level analytics. With appropriate ethical and privacy safeguards, this data can be used to refine algorithms, inform public health planning, and support regulatory evaluation of new devices^20,19^.

### Stakeholders Perspective

Patients experience both the promise and apprehension regarding adopting AI and automated devices for long-term oxygen therapy (LTOT). On the positive side, closed-loop technologies like O2matic offer dynamic titration of oxygen that can enhance safety, reduce symptom burden, and optimize autonomy in everyday life^8,23^. Intelligent, portable devices such as iPOC and edge-AI models^7,11^ support mobility and lifestyle integration. However, trust, transparency, and explainability are still concerns when decisions do not match patient expectations or are viewed as black boxes^20^. Older populations and those with diversity require inclusive design with natural interfaces and multimodal feedback^24^. Patients also have concerns regarding privacy and surveillance of data, with local processing and open consent processes^17^, as well as concern for affordability and insurance coverage, which will shape perceptions of value and uptake.

Clinicians see AI-enhanced LTOT as an opportunity to improve oxygen therapy precision and reduce manual workload, especially in ambulatory care. Systems like O2matic and iPOC have continuous oxygen titration on the basis of targets defined by clinicians, with the possibility to augment adherence and reduce clinical deterioration^7,8^. Predictive models^11^ also offer early-warning capability. However, integration with existing workflows remains a challenge, particularly when systems demand new training or are not interoperable with electronic health records^19^. Clinicians also emphasize the requirements for safety, monitoring, and transparency, particularly in the case of comorbidity, signal artifacts, or bias in underrepresented subgroups^6,16^. The ability to override or audit algorithmic decisions is essential. Furthermore, successful adoption relies on building clinician trust, supported by open legal frameworks and participatory training^17,25^.

Caregivers, including family members and home health aides, may experience significant relief from the burden of manual monitoring and adjustment when AI-based LTOT systems are employed. Automated platforms like O2matic and iPOC reduce the need for constant supervision, especially during periods of exertion or sleep, thereby improving caregiver confidence and well-being^26^. Remote monitoring features enable more flexible support, although poorly calibrated alert systems risk overwhelming caregivers with unnecessary notifications^27^. Ease of use is a critical determinant of caregiver acceptance, as they often handle equipment setup and troubleshooting, particularly when patients are physically or cognitively impaired^24^. The shift from manual operator to system supervisor necessitates new forms of training and emotional adjustment, especially as caregivers navigate trust in algorithmic decisions^20^. Their role in system procurement, insurance negotiation, and maintenance further underscores the need for affordable, reliable, and user-friendly solutions.

Regulators and policy makers play a critical role in ensuring the safe and equitable integration of AI-driven LTOT systems. As software that directly informs clinical decisions, these systems are subject to evolving guidelines for Software as a Medical Device (SaMD), emphasizing transparency, post-market surveillance, and algorithmic accountability^28,29^. Lifecycle regulation is increasingly necessary to address adaptive algorithm behavior and model drift, particularly when deployed across heterogeneous patient populations or sensor platforms^6,16^. Ensuring algorithmic equity is also a regulatory priority, as evidence grows that pulse oximeters may underperform in individuals with darker skin tones^18^. Data protection, cybersecurity, and interoperability are further key considerations under frameworks such as GDPR and HIPAA. Finally, policy makers must address gaps in reimbursement and digital access to prevent socioeconomic disparities in the uptake of intelligent oxygen technologies^27^.

### Ethical Considerations in AI-Driven Long-Term Oxygen Therapy (LTOT)

As artificial intelligence (AI) and automation are increasingly integrated into long-term oxygen therapy (LTOT) systems, a parallel emphasis must be placed on the ethical dimensions of their development, deployment, and use. These considerations are particularly salient in the context of vulnerable populations, such as older adults and individuals with chronic respiratory conditions, who often depend on LTOT for day-to-day functioning and quality of life. Ensuring that AI-driven SpO₂ monitoring and oxygen titration technologies uphold ethical standards is essential for safeguarding patient rights, equity, and trust.

#### 1. Equity and Algorithmic Fairness

One of the most pressing ethical challenges is the potential for AI models to perpetuate or exacerbate health disparities. Evidence shows that pulse oximeters—upon which most AI-driven LTOT systems rely—tend to overestimate oxygen saturation in individuals with darker skin pigmentation, increasing the risk of hypoxemia and under-treatment^16,18^. If these biased data serve as inputs for AI models without proper mitigation, downstream decisions, such as predictive oxygen dosing, may systematically disadvantage certain racial or ethnic groups.

To address this, developers and researchers must ensure that AI models are trained and validated on diverse datasets and that performance is stratified and reported by demographic variables, including skin tone, sex, and age^20^. Regulatory bodies such as the U.S. FDA now encourage inclusion of representative populations and transparency regarding performance variation across subgroups^29^. Ethically, this aligns with the principle of justice, ensuring fair access to the benefits of AI and preventing algorithmic harm to marginalized populations.

#### 2. Transparency and Explainability

Another ethical imperative is transparency—both in how AI systems function and in how their outputs are communicated to patients and clinicians. Many AI models, particularly deep learning architectures, operate as “black boxes,” offering limited visibility into the rationale behind their recommendations^20,25^. This lack of explainability can erode trust and impede informed decision-making.

In the context of LTOT, where real-time oxygen adjustments affect critical physiological processes, clinicians and patients must be able to understand the basis of algorithmic decisions. Systems should include explainable AI (XAI) components that provide interpretable outputs or rationale for changes in oxygen flow. This transparency supports autonomy by enabling users to make informed choices and challenge or override automated decisions when needed (Floridi et al., 2018).

#### 3. Autonomy and Informed Consent

Respect for patient autonomy is a foundational bioethical principle, and it is challenged when decisions are increasingly delegated to opaque AI systems. In AI-enhanced LTOT, autonomy is preserved when patients are informed of how AI components operate, what data are collected, and how decisions are made. Informed consent should be obtained not only for use of the device itself, but also for the collection and processing of continuous physiological data (Briggs & Sokol, 2021).

Moreover, patients should be given the option to opt out of fully automated oxygen regulation, either temporarily or permanently, and should retain control over key parameters when feasible. This is particularly important in situations where users perceive the algorithm’s behavior as discordant with their lived experience (e.g., breathlessness not triggering increased oxygen flow).

#### 4. Privacy, Surveillance, and Data Governance

AI-driven LTOT systems often rely on cloud connectivity and continuous data logging, which raises concerns about data privacy, surveillance, and secondary data use. Sensitive physiological and behavioral data—such as oxygenation trends, physical activity, and even sleep patterns—may be stored or transmitted, potentially exposing users to privacy breaches or non-consensual data exploitation^17,19^.

Developers must ensure compliance with legal frameworks such as the Health Insurance Portability and Accountability Act (HIPAA) in the U.S. and the General Data Protection Regulation (GDPR) in Europe. Beyond compliance, ethical data governance requires data minimization, user access to data logs, and clear policies on who can access and analyze the data^25^.

#### 5. Accountability and Liability

As AI systems gain decision-making authority, questions arise about who is accountable when adverse outcomes occur. If a patient is harmed due to an erroneous prediction or system failure, determining responsibility—whether it lies with the clinician, device manufacturer, software developer, or healthcare institution—can be legally and ethically complex^30^.

Ethical implementation requires clear delineation of responsibilities, including the establishment of human oversight mechanisms and escalation pathways. Clinicians must be empowered to override automated decisions, and patients must be informed of the chain of accountability should errors arise.

## Conclusion

This systematic review critically examined the current landscape of artificial intelligence (AI) and automated systems developed for continuous oxygen saturation (SpO₂) monitoring in the context of long-term oxygen therapy (LTOT). Across eight rigorously selected studies, we identified substantial technological progress in both AI-based SpO₂ estimation and closed-loop oxygen titration systems. These advances, while distinct in their mechanisms and maturity, collectively represent a pivotal shift from static, manual oxygen delivery toward responsive, personalized, and context-aware oxygen therapy.

AI-based approaches demonstrated exceptional technical performance, with reported mean absolute errors (MAEs) as low as 0.57% and root mean square errors (RMSEs) below 1%, even in low-perfusion or motion-prone signal conditions^5,6^. These models harness sophisticated architectures such as Gaussian process regression and deep neural networks to derive oxygenation metrics from raw photoplethysmographic (PPG) data. Moreover, studies such as Cabanas et al.^6^ highlighted the potential for algorithmic bias mitigation through skin tone–stratified analysis, addressing a critical ethical concern in pulse oximetry accuracy^16,18^. Despite these strengths, real-world validation of these models remains limited, and integration into LTOT workflows—particularly in wearable or home-based contexts—has not yet been realized.

Conversely, non-AI automated systems such as O2matic^8^ and iPOC^7^ demonstrated high clinical relevance and usability, maintaining SpO₂ within target ranges and reducing the need for clinician intervention. These closed-loop devices operate through rule-based logic, adjusting oxygen flow in real time based on continuous saturation input, and have been tested in hospital and ambulatory environments. Their success in stabilizing oxygenation during exertion or acute exacerbation illustrates their readiness for clinical deployment, though they lack the predictive personalization and adaptability offered by AI systems.

A key insight from this review is that AI and automation serve complementary roles in the evolution of LTOT. AI excels in anticipatory and adaptive modeling—tailoring oxygen delivery based on individualized patterns—while automation offers immediate, reliable titration within predefined safety parameters. Future systems should aim to integrate these strengths, creating hybrid platforms that combine real-time responsiveness with long-term learning and personalization. This direction aligns with broader healthcare trends toward precision medicine and digital chronic disease management^17^.

However, significant gaps persist. Risk of bias remains a concern, particularly in AI studies trained on homogenous or idealized datasets, where algorithm robustness under diverse, real-world conditions remains untested. Many studies lacked demographic representation, usability data, or clear reporting on algorithm transparency and explainability—factors that are essential for both clinician trust and ethical deployment^20,25^. Moreover, few systems addressed the practical challenges of LTOT in underserved settings, such as offline functionality, affordability, and ease of use for elderly or non-technologically literate users^24^.

From a stakeholder standpoint, clinicians valued the potential of automation to reduce workload and maintain physiological stability, but also raised concerns around system interpretability and liability^19^. Patients and caregivers welcomed the promise of reduced burden and improved oxygen control, yet highlighted the need for transparency, control, and data privacy^17,26^.

Regulators, meanwhile, are challenged to adapt evolving frameworks to ensure safety, equity, and accountability in these emerging systems^29,30^.

In conclusion, the convergence of AI and automation is reshaping the possibilities of long-term oxygen therapy. While closed-loop systems have already proven feasible and clinically beneficial, the next frontier lies in responsibly translating AI models into intelligent, patient-centered LTOT systems that learn, adapt, and support equitable, high-quality respiratory care. Achieving this will require interdisciplinary collaboration across clinicians, engineers, ethicists, and regulators grounded in rigorous science, inclusive design, and a commitment to both innovation and safety.

## Data Availability

All data produced in the present study are available upon reasonable request to the authors

## Abbreviations

AI: Artificial Intelligence — Computer systems that mimic human intelligence to perform tasks such as learning, decision-making, and pattern recognition.
LTOT: Long-Term Oxygen Therapy — A chronic treatment involving the use of supplemental oxygen for extended periods (usually ≥15 hours/day), primarily in patients with chronic hypoxemia.
SpO₂: Peripheral Capillary Oxygen Saturation — A non-invasive measurement of the amount of oxygen-bound hemoglobin in the blood, expressed as a percentage.
PPG: Photoplethysmography — An optical technique used to detect blood volume changes in the microvascular bed of tissue, commonly used in pulse oximeters.
MAE: Mean Absolute Error — A metric used to measure the average magnitude of prediction errors in a model, without considering direction.
RMSE: Root Mean Square Error — A statistical measure of the differences between predicted values and observed values; sensitive to large errors.
FDA: Food and Drug Administration — The U.S. federal agency responsible for regulating medical devices, drugs, and diagnostics.
COPD: Chronic Obstructive Pulmonary Disease — A progressive respiratory disease characterized by airflow limitation, often treated with LTOT.
SaMD: Software as a Medical Device — Software intended for medical use that performs functions without being part of a hardware medical device.
6MWT: Six-Minute Walk Test — A clinical test used to assess functional exercise capacity, often used to determine LTOT eligibility.
iPOC: Intelligent Portable Oxygen Concentrator — A device that automatically adjusts oxygen delivery based on patient activity and real-time monitoring.
GMLP: Good Machine Learning Practices — FDA-proposed standards to ensure safety, efficacy, and accountability in AI/ML medical software.
XAI: Explainable Artificial Intelligence — A branch of AI focused on making model decision-making processes transparent and understandable to users.
EHR: Electronic Health Record — A digital version of a patient’s paper chart, containing medical history, diagnoses, medications, and treatment plans.
ABG: Arterial Blood Gas — A test that measures oxygen and carbon dioxide levels in arterial blood, used to validate SpO₂ accuracy.
MCAR /MAR: Missing Completely At Random / Missing At Random — Statistical assumptions about the mechanism of missing data in datasets.
HIPAA: Health Insurance Portability and Accountability Act — U.S. legislation that provides data privacy and security provisions for safeguarding medical information.
GDPR: General Data Protection Regulation — A legal framework that sets guidelines for the collection and processing of personal information from individuals in the EU.
SUS: System Usability Scale — A standardized questionnaire used to evaluate the usability of software or devices.
IMDRF: International Medical Device Regulators Forum — A global coalition that develops harmonized regulatory approaches for medical devices.
PRISMA: Preferred Reporting Items for Systematic Reviews and Meta-Analyses — An evidence-based set of guidelines for reporting systematic reviews.
CONSORT-AI / DECIDE-AI: Reporting extensions to the CONSORT guidelines — Adapted to trials evaluating AI interventions; emphasize transparency and reproducibility.

## Supplementary Appendix A: Inclusion/Exclusion Criteria and Search Strategies by Database

### Inclusion Criteria

Population:

- ● Clinical settings (e.g., ICU, peri-operative care) or mixed cohorts (healthy subjects + patients).

Intervention:

- ● AI-driven or Automated SpO₂ monitoring addressing LTOT-relevant technical challenges, such as:

○ Motion artifact correction
○ Low perfusion or weak signal handling.
○ Skin tone bias mitigation.
○ Long-term signal stability

Study Design:

● Peer-reviewed, conference committe-reviewed, open-access articles (no paywalls).
● English language.
● Published between 2000–2025. Outcomes:
● Technical validation of methods transferable to LTOT
● Robust preprocessing techniques

### Exclusion Criteria

Population:

● Studies focused on acute/non-chronic conditions Intervention:
● Non-AI or non-automated irrelevant to LTOT challenges (e.g., pulse rate estimation without SpO₂ focus).

Study Type:

● Conference abstracts, theses, preprints, or non-peer-reviewed articles.
● Paywalled articles or studies requiring institutional login.
● Non-English studies. Scope:
● Studies lacking explicit relevance to LTOT monitoring (e.g., AI for ECG analysis).

### 1. Included Studies

#### 1.1 AI-Based Systems (5 studies)

1. Cabanas et al., 2024 (MDPI) *Evaluating AI Methods for Pulse Oximetry: Performance, Clinical Accuracy, and Comprehensive Bias Analysis*
2. Shuzan et al., 2023 (Found through reference) *Machine Learning-Based Respiration Rate and Blood Oxygen Saturation Estimation Using Photoplethysmogram Signals*
3. Argüello-Prada & Castillo García, 2024 (Found through reference) *Machine Learning Applied to Reference Signal-Less Detection of Motion Artifacts in Photoplethysmographic Signals: A Review*
4. Pascual-Saldaña et al., 2024 (PubMed) *Innovative Predictive Approach towards a Personalized Oxygen Dosing System*
5. Pascual et al., 2023 (AI4Health prototype, IEEE) *Analyzing Distinct Neural Network Models for Oxygen Saturation Prediction Towards a Personalized COPD Management*

#### 1.2 Automated (Non-AI) Oxygen Delivery Systems (3 studies)

1. Sanchez-Morillo et al., 2020 (PubMed) *Automated Home Oxygen Delivery for Patients with COPD and Respiratory Failure: A New Approach*
2. Hansen et al., 2018 (PubMed) *Automated Oxygen Control with O2matic® During Admission with Exacerbation of COPD*
3. Cirio & Nava, 2011 (PubMed) *Pilot Study of a New Device to Titrate Oxygen Flow in Hypoxic Patients on Long-Term Oxygen Therapy*

### 2. PubMed Search Strategy

2000–2025 | English

**Search Query:**

(("oxygen therapy" OR "long-term oxygen therapy" OR LTOT OR "oxygen titration" OR "oxygen delivery" OR "oxygen therapy")

AND ("artificial intelligence" OR "machine learning" OR "deep learning"

OR "automated system" OR "closed-loop" OR automation OR "smart device" OR "predictive model" OR "digital health" OR "closed-loop")

AND ("SpO2" OR "oxygen saturation" OR "pulse oximetry" OR photoplethysmography OR PPG OR oximeter))

**Total Found:** 111

**Title & Abstract Screening:** 111

**Full-Text Review:** 15

1. *New Perspectives in Oxygen Therapy Titration: Is Automatic Titration the Future?*
2. *Innovative Predictive Approach towards a Personalized Oxygen Dosing System*
3. *Automated O*₂ *Titration Alone or With High-Flow Nasal Cannula During Walking Exercise in Chronic Lung Diseases*
4. *Automated Home Oxygen Delivery for Patients with COPD and Respiratory Failure: A New Approach*
5. *Automated Oxygen Control with O2matic® During Admission with Exacerbation of COPD*
6. *Pilot Study of a New Device to Titrate Oxygen Flow in Hypoxic Patients on Long-Term Oxygen Therapy*
7. *Machine Learning Prediction for Supplemental Oxygen Requirement in Patients with COVID-19*
8. *Home Oxygen Therapy: Re-thinking the Role of Devices*
9. *Usability of a Continuous Oxygen Saturation Device for Home Telemonitoring*
10. *The Experience of Automated Home Oxygen Therapy for Patients With COPD – A Qualitative Study*
11. *Automatic versus Manual Oxygen Titration in Patients Requiring Supplemental Oxygen in the Hospital: A Systematic Review and Meta-Analysis*
12. *Innovative Predictive Approach towards a Personalized Oxygen Dosing System*
13. *Recent Advances in Wearable Sensors and Data Analytics for Continuous Monitoring and Analysis of Biomarkers and Symptoms Related to COVID-19*
14. *Evaluation of a Novel Ear Pulse Oximeter: Towards Automated Oxygen Titration in Eyeglass Frames*
15. *Cost-effectiveness of FreeO*₂ *in Patients with COPD Hospitalized for Acute Exacerbations: Analysis of a Pilot Study in Quebec*

- **Included:** 4
  1. Innovative Predictive Approach towards a Personalized Oxygen Dosing System
  2. Automated Home Oxygen Delivery for Patients with COPD and Respiratory Failure: A New Approach
  3. Automated Oxygen Control with O2matic® During Admission with Exacerbation of COPD
  4. Pilot Study of a New Device to Titrate Oxygen Flow in Hypoxic Patients on Long-Term Oxygen Therapy

### 3. IEEE Xplore

2000–2025 | English

Search Query:

(("oxygen therapy" OR "long-term oxygen therapy" OR LTOT

OR "oxygen titration" OR "oxygen delivery" OR "oxygen therapy")

AND ("artificial intelligence" OR "machine learning" OR "deep learning"

OR "automated system" OR "closed-loop" OR automation OR "smart device"

OR "predictive model" OR "digital health" OR "closed-loop")

AND ("SpO2" OR "oxygen saturation" OR "pulse oximetry"OR photoplethysmography OR PPG OR oximeter))

**Total Found:** 23

**Title & Abstract Screening:** 3 selected

- Analyzing Distinct Neural Network Models for Oxygen Saturation Prediction Towards a Personalized COPD Management
- Smart Ventilation Bag with Adjustable Oxygen Range and Automated Pressure Regulation
- Influence-Based Nano Fuzzy Swarm Oxygen Deficiency Detection and Therapy

**Final Selected:** 1

- Analyzing Distinct Neural Network Models for Oxygen Saturation Prediction Towards a Personalized COPD Management

### 4. Springer Link

2000–2025 | English | Open Access

Search Query:

(("chronic obstructive pulmonary disease" OR COPD OR "chronic lung disease")

AND ("digital health" OR "eHealth" OR "remote monitoring" OR "telemedicine")

AND ("artificial intelligence" OR "machine learning" OR "deep learning"

OR "predictive model" OR "automated system" OR "closed-loop"

OR automation OR "smart device")

AND ("oxygen therapy" OR "long-term oxygen therapy" OR LTOT

OR "oxygen titration" OR "oxygen delivery")

AND ("SpO2" OR "oxygen saturation" OR "pulse oximetry"

OR photoplethysmography OR PPG OR oximeter))

**Articles Found:** 50

**Full-Text Screening:** 5

1. *Applications of digital health technologies and artificial intelligence algorithms in COPD: systematic review*
2. *Machine-learning based feature selection for a non-invasive breathing change detection*
3. *Continuous remote monitoring of COPD patients—justification and explanation of the requirements and a survey of the available technologies*
4. *A systematic review of the impacts of remote patient monitoring (RPM) interventions on safety, adherence, quality of life, and cost-related outcomes*
5. *Feasibility and acceptability of remotely monitoring spirometry and pulse oximetry as part of interstitial lung disease clinical care*

**Final Inclusion:** None

### 5. MDPI Searches

*(Multiple searches were performed due to the MDPI advanced search not supporting free-text queries. Where applicable, screenshots or figures were captured to document search parameters and filters used.)*

**Search #1:** “Oxygen Delivery” AND “LTOT” AND “Automated”

**Records Screened:** 2

**Included:** 1

*Automated Home Oxygen Delivery for Patients with COPD and Respiratory Failure: A New Approach*

**Search #2:** “Oxygen Delivery” AND “LTOT” AND “Artificial Intelligence”

**Records Screened:** 0

**Included:** 0

**Search #3:** “Oxygen Delivery” AND “LTOT” AND “Machine Learning”

**Records Screened:** 0

**Included:** 0

**Search #4:** “COPD” AND “LTOT” AND “Machine Learning”

**Records Screened:** 4

**Included:** 1

*Innovative Predictive Approach towards a Personalized Oxygen Dosing System*

**Search #5:** “SpO2” AND “Artificial Intelligence” OR “Machine Learning”

**Records Screened:** 5

**Included:** 1

*Evaluating AI Methods for Pulse Oximetry: Performance, Clinical Accuracy, and Comprehensive Bias Analysis*

### 6. ACMI (Association for Computing Machinery)

Search Query:

(“Machine Learning” OR “Artificial Intelligence”) AND (SpO₂ OR “oxygen delivery”)

**Total Found:** 731

**Title & Abstract Screening:** 15

1. *A Machine-Learning-Based Prediction Method for Easy COPD Classification Based on Pulse Oximetry (Abineza et al., 2022)*
2. *A Survey on Pre-Training Requirements for Deep Learning Models to Detect Obstructive Sleep Apnea*
3. *Classification of the Sleep-Wake State Through the Development of a Deep Learning Model*
4. *Deployment of Artificial Intelligence Models for Sleep Apnea Recognition in the Sleep Laboratory*
5. *Scale-Based Entropy Measures and Deep Learning Methods for Analyzing Cardiorespiratory Control in COVID-19*
6. *Predicting Mixed Venous Oxygen Saturation (SvO*₂*) Impairment in COPD Patients Using Clinical-CT Radiomics*
7. *Smartphone-Based SpO*₂ *Measurement Using Near-IR and Chromophore Compensation*
8. *Toward Sleep Apnea Detection with Lightweight Multi-Scaled Fusion Network*
9. *Hybridization of Soft-Computing Algorithms with Neural Network for Obstructive Sleep Apnea Prediction*
10. *Screening of Finger Pulse Oximeter Pre-Calibration Function Test with Machine Learning*
11. *Deep Learning Approach to Estimate SpO*₂ *from PPG Signals*
12. *Non-Contact PPG Signal and Heart Rate Estimation with Multi-Hierarchical Convolutional Network*
13. *Comparison of SFS and mRMR for Oximetry Feature Selection in Obstructive Sleep Apnea Detection*
14. *Deep Learning–Based PPG Quality Assessment for Heart Rate and Variability*
15. *Automated Medical Oxygen Regulator Using Blood Oxygen Saturation Level*

**Full-Text Screening:** 5

16. A Machine-Learning-Based Prediction Method for Easy COPD Classification Based on Pulse Oximetry (Abineza et al., 2022)
17. Smartphone-Based SpO₂ Measurement Using Near-IR and Chromophore Compensation (Bui et al., 2017/2020)
18. Screening of Finger Pulse Oximeter Pre-Calibration Function Test with Machine Learning (Khairunnisa et al., 2024)
19. Deep Learning Approach to Estimate SpO₂ from PPG Signals (Koteska et al., 2023)
20. Automated Medical Oxygen Regulator Using Blood Oxygen Saturation Level (Linsangan et al., 2023)

**All Rejected** (paywall restrictions)

## Supplementary Appendix B: Risk of Bias Analysis

<u>R code:</u>

library(dplyr) library(robvis)

levels_robvis ← c("Low", "Moderate", "Serious", "Critical", "No information") risk_of_bias_mod ← data.frame(

Study = c(

"Cabanas et al. (2024)",

"Shuzan et al. (2023)",

"Argüello-Prada & Castillo (2024)",

"Pascual-Saldaña et al. (2024)",

"AI4Health Prototype (2023)",

"Sanchez-Morillo et al. (2020)",

"Hansen et al. (2018)",

"Cirio & Nava (2011)"

),

Domain1 = c("Moderate", "Moderate", "Moderate", "Moderate", "Serious", "Moderate", "Moderate", "Serious"),

Domain2 = c("Moderate", "Serious", "Serious", "Moderate", "Moderate", "Moderate", "Moderate", "Moderate"),

Domain3 = c("Low", "Low", "Moderate", "Low", "Serious", "Low", "Low", "Low"),

Domain4 = c("Low", "Moderate", "Moderate", "Moderate", "Serious", "Low", "Low", "Low"),

Domain5 = c("Low", "Moderate", "Moderate", "Moderate", "Moderate", "Low", "Low", "Low"),

Domain6 = c("Low", "Moderate", "Moderate", "Moderate", "Serious", "Moderate", "Moderate", "Serious"),

Domain7 = c("Low", "Moderate", "Moderate", "Moderate", "Serious", "Moderate", "Moderate", "Serious"), Overall = c("Moderate", "Serious", "Serious", "Moderate", "Serious", "Moderate", "Moderate", "Serious"),

Weight = c(1, 1, 1, 1, 1, 1, 1, 1),

stringsAsFactors = FALSE

)

risk_of_bias_mod ← risk_of_bias_mod %>%

mutate(across(starts_with("Domain"), ∼ factor(., levels = levels_robvis, ordered = TRUE))) %>%

mutate(Overall = factor(Overall, levels = levels_robvis, ordered = TRUE))

str(risk_of_bias_mod)

rob_summary(data = risk_of_bias_mod, tool = "ROBINS-I", weighted = TRUE)

rob_traffic_light(data = risk_of_bias_mod, tool = "ROBINS-I")

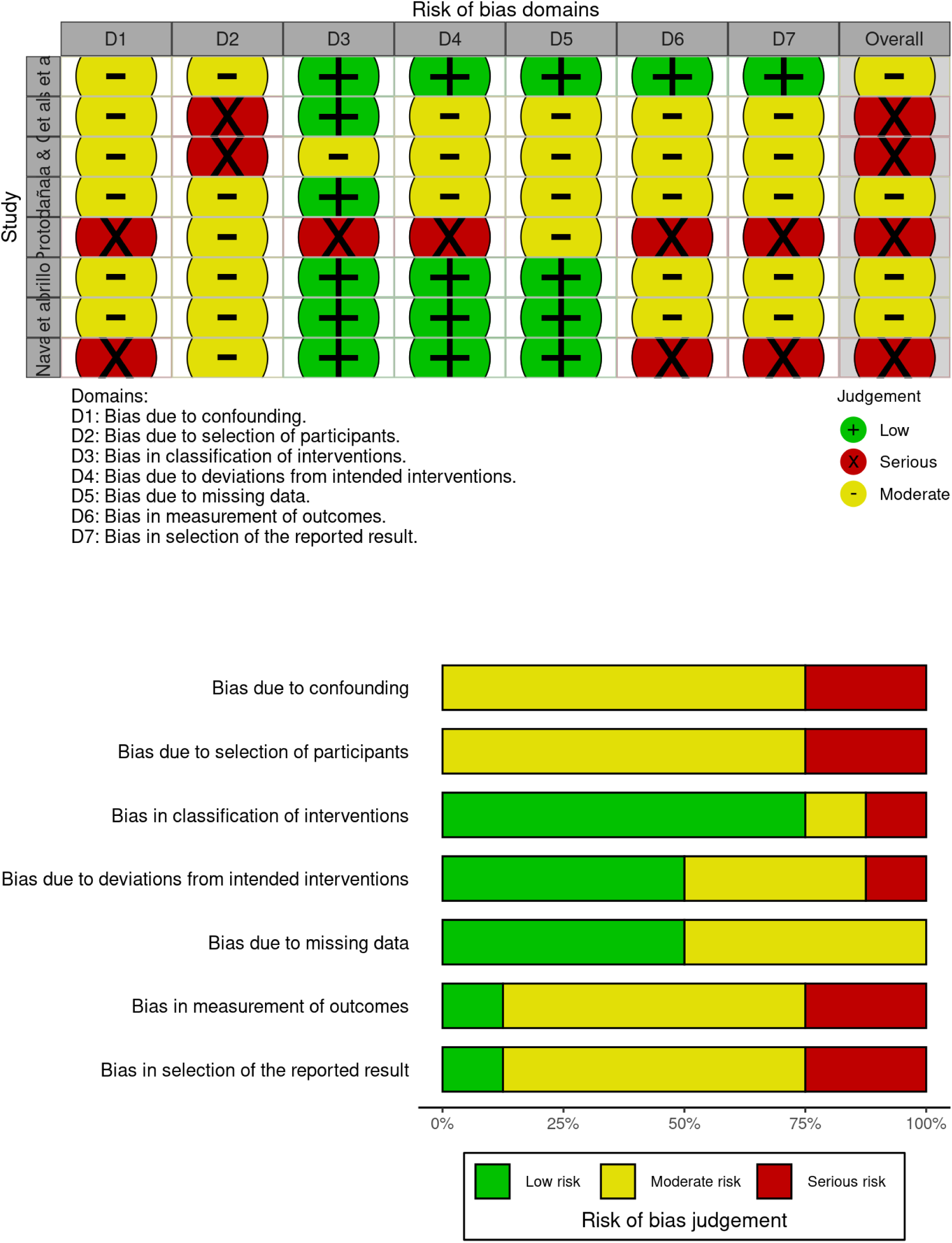

## References

1. World Health Organization. Global Health Estimates: Life expectancy and leading causes of death and disability.2020. https://www.who.int/data/gho/data/themes/mortality-and-global-health-estimates

2. Nocturnal Oxygen Therapy Trial Group. Continuous or Nocturnal Oxygen Therapy in Hypoxemic Chronic Obstructive Lung Disease. Ann Intern Med. 1980;93(3):391–398. doi:10.7326/0003-4819-93-3-391

3. Medical Research Council Working Party. Long term domiciliary oxygen therapy in chronic hypoxic cor pulmonale complicating chronic bronchitis and emphysema. Report of the Medical Research Council Working Party. Lancet. 1981;1(8222):681–686.

4. Global Initiative for Chronic Obstructive Lung Disease (GOLD). Global Strategy for Prevention, Diagnosis, and Management of COPD: 2024 Report. Published 2024. https://goldcopd.org/2024-gold-report/

5. Shuzan MNI, Chowdhury MH, Chowdhury MEH, Murugappan M, Hoque Bhuiyan E, Arslane Ayari M, Khandakar A. Machine Learning-Based Respiration Rate and Blood Oxygen Saturation Estimation Using Photoplethysmogram Signals. Bioengineering. 2023; 10(2):167. 10.3390/bioengineering10020167

6. Cabanas AM, Sáez N, Collao-Caiconte PO, Martín-Escudero P, Pagán J, Jiménez-Herranz E, Ayala JL. Evaluating AI Methods for Pulse Oximetry: Performance, Clinical Accuracy, and Comprehensive Bias Analysis. Bioengineering. 2024; 11(11):1061. 10.3390/bioengineering11111061

7. Sanchez-Morillo D, Muñoz-Zara P, Lara-Doña A, Leon-Jimenez A. Automated Home Oxygen Delivery for Patients with COPD and Respiratory Failure: A New Approach. Sensors. 2020; 20(4):1178. 10.3390/s20041178

8. Hansen EF, Hove JD, Bech CS, Jensen JS, Kallemose T, Vestbo J. Automated oxygen control with O2matic® during admission with exacerbation of COPD. Int J Chron Obstruct Pulmon Dis. 2018;13:3997–4003. Published 2018 Dec 14. doi:10.2147/COPD.S183762

9. Moher D, Liberati A, Tetzlaff J, Altman DG; PRISMA Group. Preferred reporting items for systematic reviews and meta-analyses: the PRISMA statement. PLoS Med. 2009;6(7):e1000097. doi:10.1371/journal.pmed.1000097

10. Argüello-Prada EJ, Castillo García JF. Machine Learning Applied to Reference Signal-Less Detection of Motion Artifacts in Photoplethysmographic Signals: A Review. Sensors. 2024; 24(22):7193. 10.3390/s24227193

11. Pascual-Saldaña H, Masip-Bruin X, Asensio A, Alonso A, Blanco I. Innovative Predictive Approach towards a Personalized Oxygen Dosing System. Sensors. 2024; 24(3):764. 10.3390/s24030764

12. Liu X, Cruz Rivera S, Moher D, et al. Reporting guidelines for clinical trial reports for interventions involving artificial intelligence: the CONSORT-AI extension. Nat Med. 2020;26:1364–1374. doi:10.1038/s41591-020-1034-x

13. Vasey B, Nagendran M, Campbell B, et al. Reporting guideline for the early stage clinical evaluation of decision support systems driven by artificial intelligence: DECIDE-AI. BMJ. 2022;377:e070904. Published 2022 May 18. doi:10.1136/bmj-2022-070904

14. Pascual H, Masip-Bruin X, Alonso A, Blanco I. Analyzing distinct neural network models for oxygen saturation prediction towards a personalized COPD management. In: 2023 IEEE 19th International Conference on e-Science (e-Science). IEEE; 2023:1–8. https://api.semanticscholar.org/CorpusID:262980603

15. Cirio S, Nava S. Pilot study of a new device to titrate oxygen flow in hypoxic patients on long-term oxygen therapy. Respir Care. 2011;56(4):429–434. doi:10.4187/respcare.00983

16. Sjoding MW, Dickson RP, Iwashyna TJ, Gay SE, Valley TS. Racial bias in pulse oximetry measurement. N Engl J Med. 2020;383(25):2477–2478. doi:10.1056/NEJMc2029240

17. Topol EJ. High-performance medicine: the convergence of human and artificial intelligence. Nat Med. 2019;25:44–56. doi:10.1038/s41591-018-0300-7

18. Fawzy A, Wu TD, Wang K, et al. Racial and Ethnic Discrepancy in Pulse Oximetry and Delayed Identification of Treatment Eligibility Among Patients With COVID-19 [published correction appears in JAMA Intern Med. 2022 Oct 1;182(10):1108. doi: 10.1001/jamainternmed.2022.3817.]. JAMA Intern Med. 2022;182(7):730–738. doi:10.1001/jamainternmed.2022.1906

19. Shortliffe EH, Sepúlveda MJ. Clinical decision support in the era of artificial intelligence. JAMA. 2018;320(21):2199–2200. doi:10.1001/jama.2018.17163

20. London AJ. Artificial intelligence and black-box medical decisions: accuracy versus explainability. Hastings Cent Rep. 2019;49(1):15–21. doi:10.1002/hast.973

21. Hardinge M, Annandale J, Bourne S, et al. British Thoracic Society guidelines for home oxygen use in adults. Thorax. 2015;70 Suppl 1:i1–i43. doi:10.1136/thoraxjnl-2015-206865

22. Chan ED, Chan MM, Chan MM. Pulse oximetry: understanding its basic principles facilitates appreciation of its limitations. Respir Med. 2013;107(6):789–799. doi:10.1016/j.rmed.2013.02.004

23. Plant PK, Owen JL, Elliott MW. Early use of non-invasive ventilation for acute exacerbations of chronic obstructive pulmonary disease on general respiratory wards: a multicentre randomised controlled trial. Lancet. 2000;355(9219):1931-1935. doi:10.1016/S0140-6736(00)02323-0

24. Vaportzis E, Clausen MG, Gow AJ. Older Adults Perceptions of Technology and Barriers to Interacting with Tablet Computers: A Focus Group Study. Front Psychol. 2017;8:1687. doi:10.3389/fpsyg.2017.01687

25. Morley J, Machado CCV, Burr C, et al. The ethics of AI in health care: A mapping review. Soc Sci Med. 2020;260:113172. doi:10.1016/j.socscimed.2020.113172

26. Wittenberg E, Goldsmith JV, Chen C, Prince-Paul M, Johnson RR. Opportunities to improve COVID-19 provider communication resources: A systematic review. Patient Educ Couns. 2021;104(3):438–451. doi:10.1016/j.pec.2020.12.031

27. Kelly CJ, Karthikesalingam A, Suleyman M, et al. Key challenges for delivering clinical impact with artificial intelligence. BMC Med. 2019;17:195. doi:10.1186/s12916-019-1426-2

28. International Medical Device Regulators Forum (IMDRF). Medical Devices: Post-Market Surveillance: National Competent Authority Report Exchange Criteria and Report Form. National Competent Authority Report Working Group. September 21, 2017. https://www.imdrf.org/sites/default/files/docs/imdrf/final/technical/imdrf-tech-170921-pms-ncar-n14-r2.pdf

29. U.S. Food and Drug Administration (FDA). Artificial Intelligence/Machine Learning (AI/ML)-Based Software as a Medical Device (SaMD) Action Plan. January 2021. https://www.fda.gov/media/145022/download

30. Gerke S, Minssen T, Cohen G. Ethical and legal challenges of artificial intelligence-driven healthcare. In: Bohr A, Memarzadeh K, eds. Artificial Intelligence in Healthcare. Academic Press; 2020:295–336. doi:10.1016/B978-0-12-818438-7.00012-5

